# Coagulopathy in patients with Coronavirus Disease 2019 (COVID-19): A systematic review and meta-analysis

**DOI:** 10.1101/2020.07.15.20154138

**Authors:** Xiaolin Zhang, Xue Yang, Hongmei Jiao, Xinmin Liu

## Abstract

Patients with COVID-19 frequently manifest coagulation abnormalities and thrombotic events. In this meta-analysis, we aimed to explore the role of coagulopathy on the severity differences in patients with COVID-19. We conducted systematic literature search via Pubmed, Embase, Cochrane, WanFang Database, CNKI, and medRxiv from December 1, 2019 to May 1, 2020, to identify all original studies that reports on coagulation parameters (D-dimer, PLT, PT, APTT, and FIB) during COVID-19 infection. Thereafter, we compared the coagulation parameters between less severe and more severe cases. All Statistical analyses were performed via Stata14.0 software. A total of 3,952 confirmed COVID-19 infected patients were included from 25 studies. Patients with severe COVID-19 infection exhibited significantly higher levels of D-dimer, PT, and FIB (SMD 0.83, 95% CI: 0.70-0.97, I^2^ 56.9%; SMD 0.39, 95% CI: 0.14-0.64, I^2^ 77.9%; SMD 0.35, 95% CI: 0.17-0.53, I^2^42.4% respectively). However, difference in PLT and APTT levels between less severe and more severe patients was not statistically significant (SMD-0.26, 95% CI:-0.56-0.05, I^2^ 82.2%; SMD-0.14,95% CI: −0.45-0.18, I^2^ 75.5% respectively) This meta-analysis revealed coagulopathy is associated with the severity of COVID-19. Notably, D-dimer, PT, and FIB are the dominant parameters that should be considered in evaluating coagulopathy in COVID-19 patients.

## Introduction

Coronavirus Disease 2019 (COVID-19) is a viral respiratory infection caused by the 2019 novel coronavirus. A total of 5,061,476 confirmed cases with 331,475 deaths have been reported globally since its outbreak on May 22, 2020 [1]. Thus, the World Health Organization declared the disease a global pandemic. COVID-19 is the third most lethal zoonotic coronavirus disease after SARS and MERS that occurred in the last two decades [2], it has however caused many deaths than SARS or MERS [3]. Due to the extremely high mortality rate of COVID-19, there is an urgent need to identify clinical features associated with its progression.

Notably, coagulopathy and thrombotic events are prominent in some COVID-19 patients. Following the previous researches in China, highly prevalent coagulopathy characterized by elevated PT, APTT, and D-dimer was reported [4]. Besides, several studies reported that elevated D-dimer levels were highly associated with in-hospital mortality [5, 6]. Other than lung injuries, recent autopsy reports from the United States confirmed that severe COVID-19 patients exhibit a hypercoagulable state evidenced by thrombotic events in the lung, kidney, and possibly in the heart and other organs [7]. However, whether coagulopathy is associated with the severity of COVID-19 and the features of coagulation dysfunction remains unclear. Therefore, we conducted a systematic review and meta-analysis by incorporating both English and Chinese published literature to explore the coagulation dysfunction on the severity of COVID-19 progression.

## Results

### Research selection and quality assessment

The detailed literature search steps are highlighted in the flow diagram (Figure 1). We identified 1,967 records using different search strategies in the six databases. After eliminating the duplicate records, 1,132 articles were obtained. Besides, an additional 776 articles were excluded because they were not original studies or irrelevant to our meta-analysis. Full texts of the remaining 356 articles were assessed for eligibility, out of which 322 were eliminated for meeting the exclusion criteria. A total of 34 articles were included in the quality evaluation from which 9 were excluded due to low quality (NOS<5). Eventually, we included 25 articles (1 in Chinese and 24 in English) for analyses [9-33]. The overall quality of available literature was moderate or high with NOS scores ranging from 5-7. The quality evaluation of the 34 articles is shown in Supplementary Table 2.

**Figure 1.**
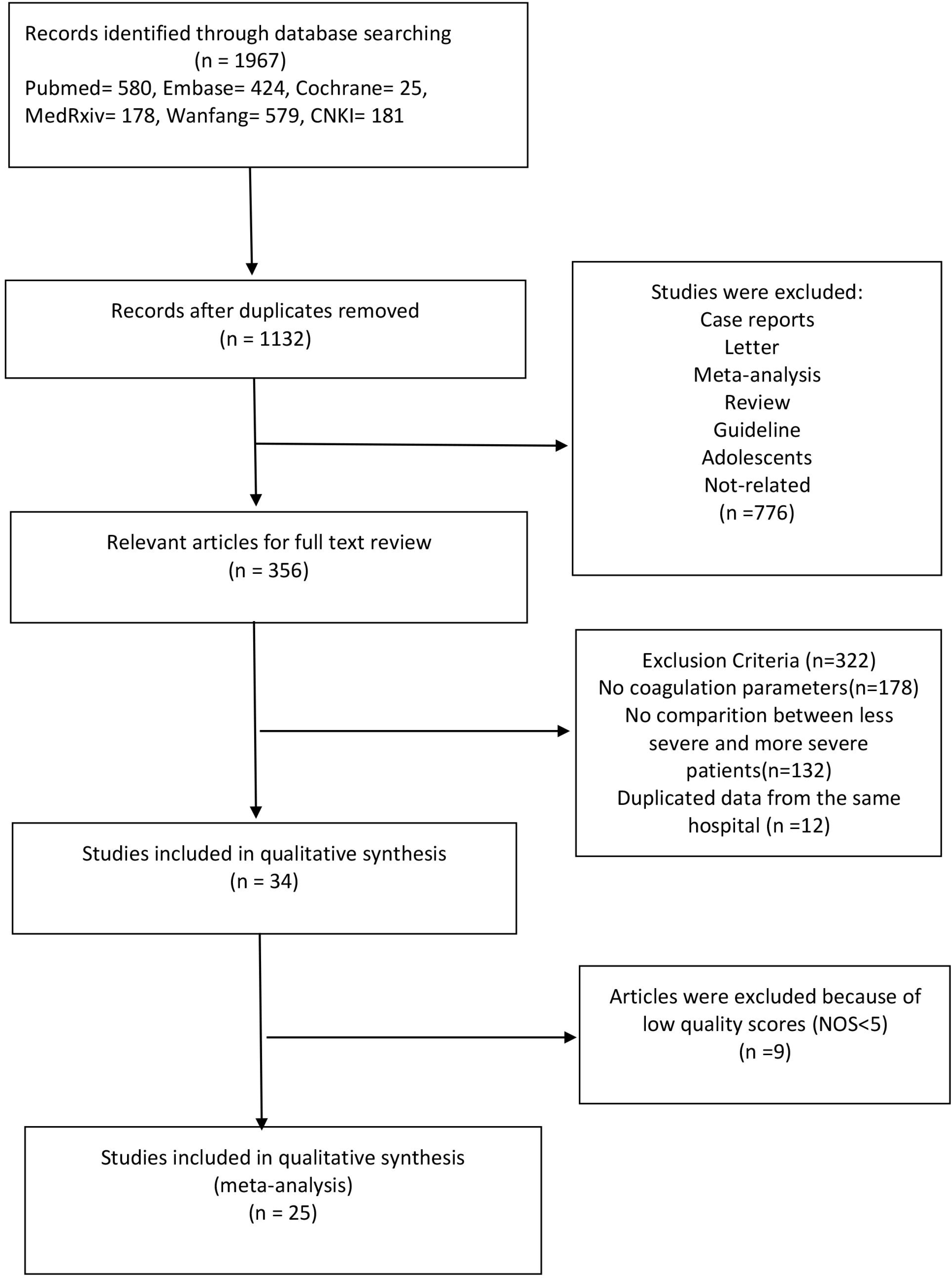
Systematic searches of the literature.

### Characteristics of the included studies

A total of 3,952 patients in the 25 included studies provided data describing coagulation parameters in the less severe and more severe groups. Notably, 23 studies had been conducted in China (11 from Wuhan and 12 from other cities), 1 from Mexico, and 1 from the USA. All the studies were retrospective observational studies and sample size varied from 21 to 577. The median age ranged between 46 and 60 years and the proportion of male patients ranged between 35.3% and 81.0%. Twenty studies followed clinical guidelines (including trial version 4, trial version 5, trial version 7, 6^th^ edition guidelines, 7^th^ edition guidelines, and WHO interim guideline) to judge on the disease severity. However, two studies considered whether patients experienced ARDS, and one study considered whether patients experienced ICU care to judge the severity of the disease. The patient characteristics and demographic data for the included studies are shown in Table 1.

**Table 1.**
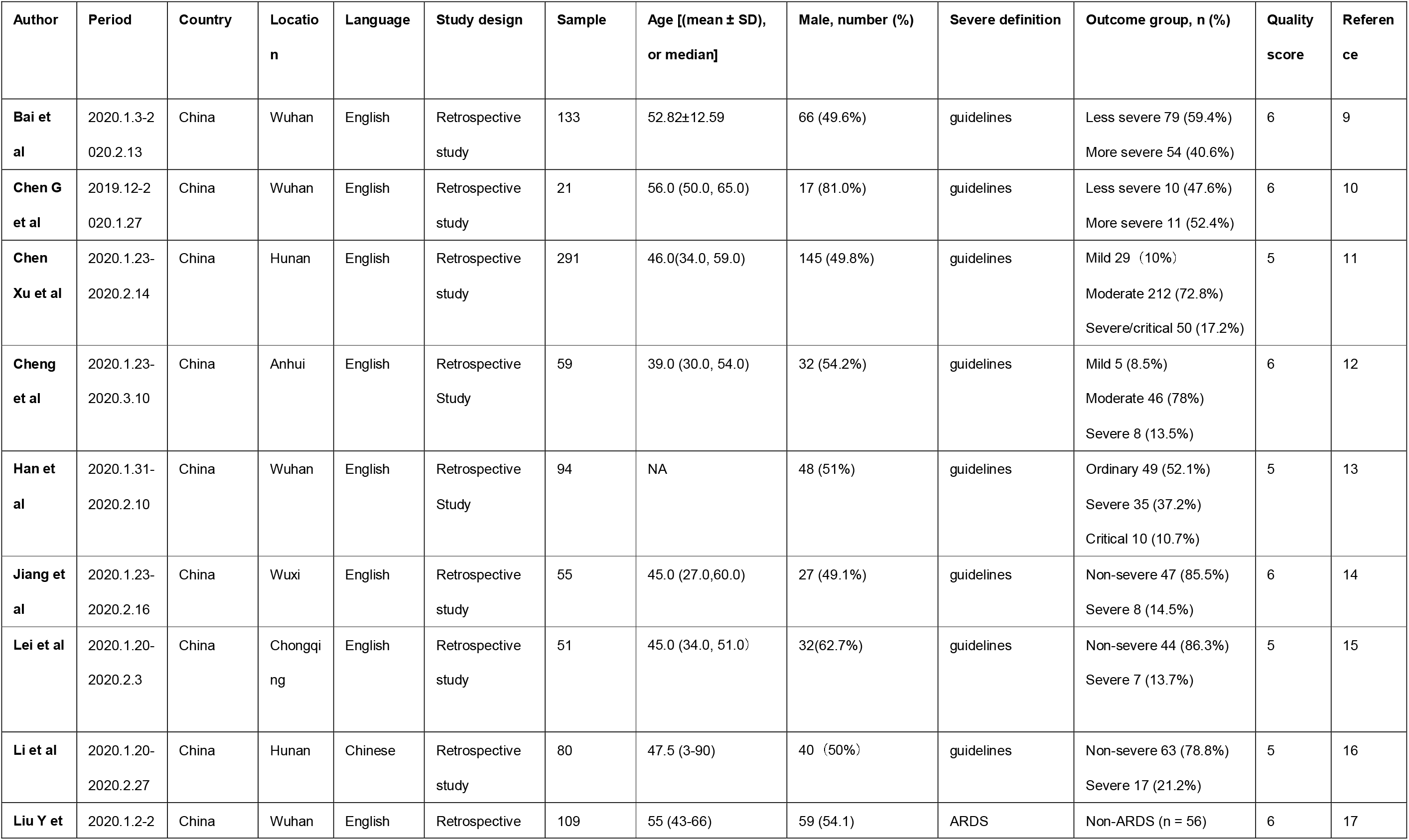
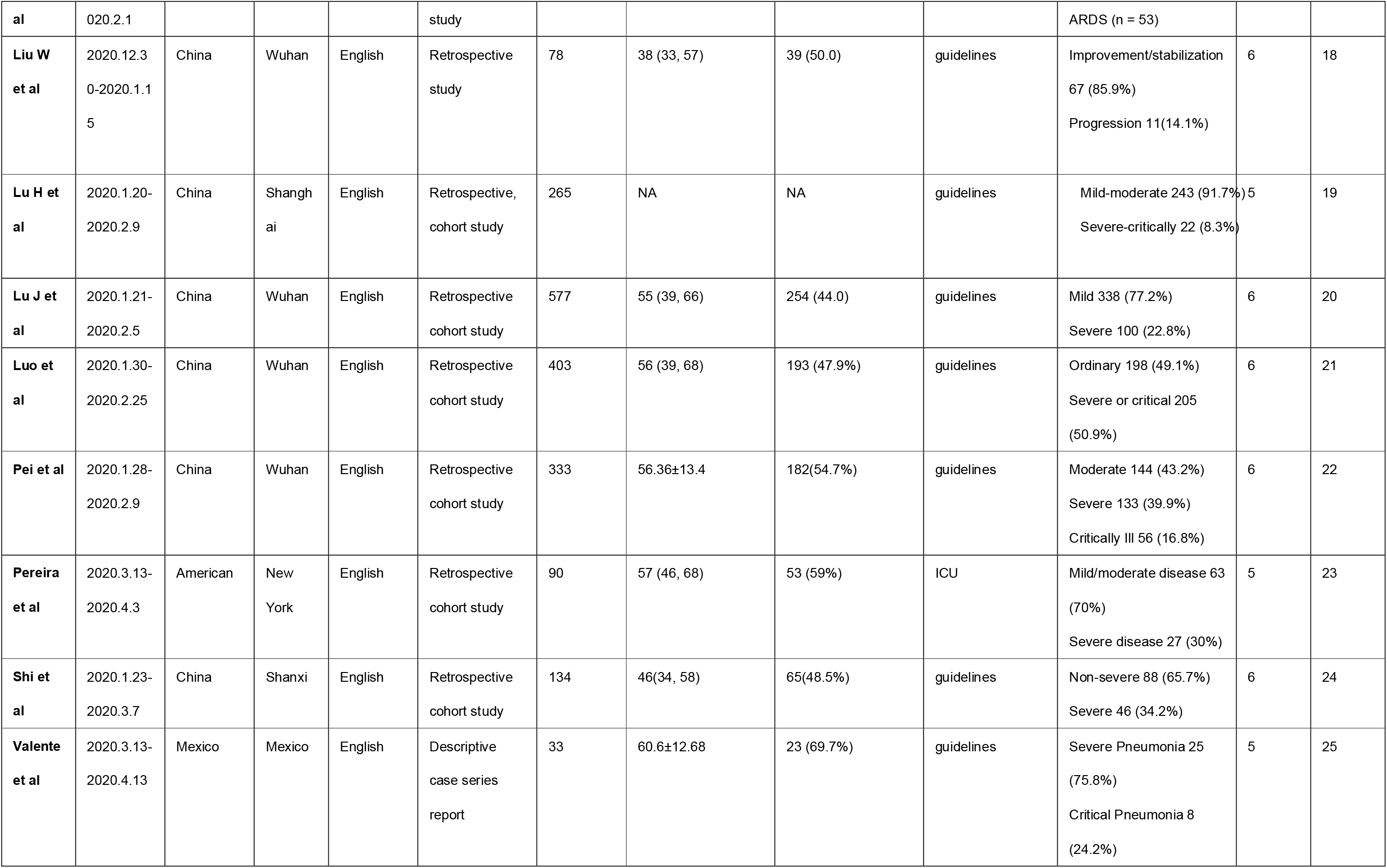
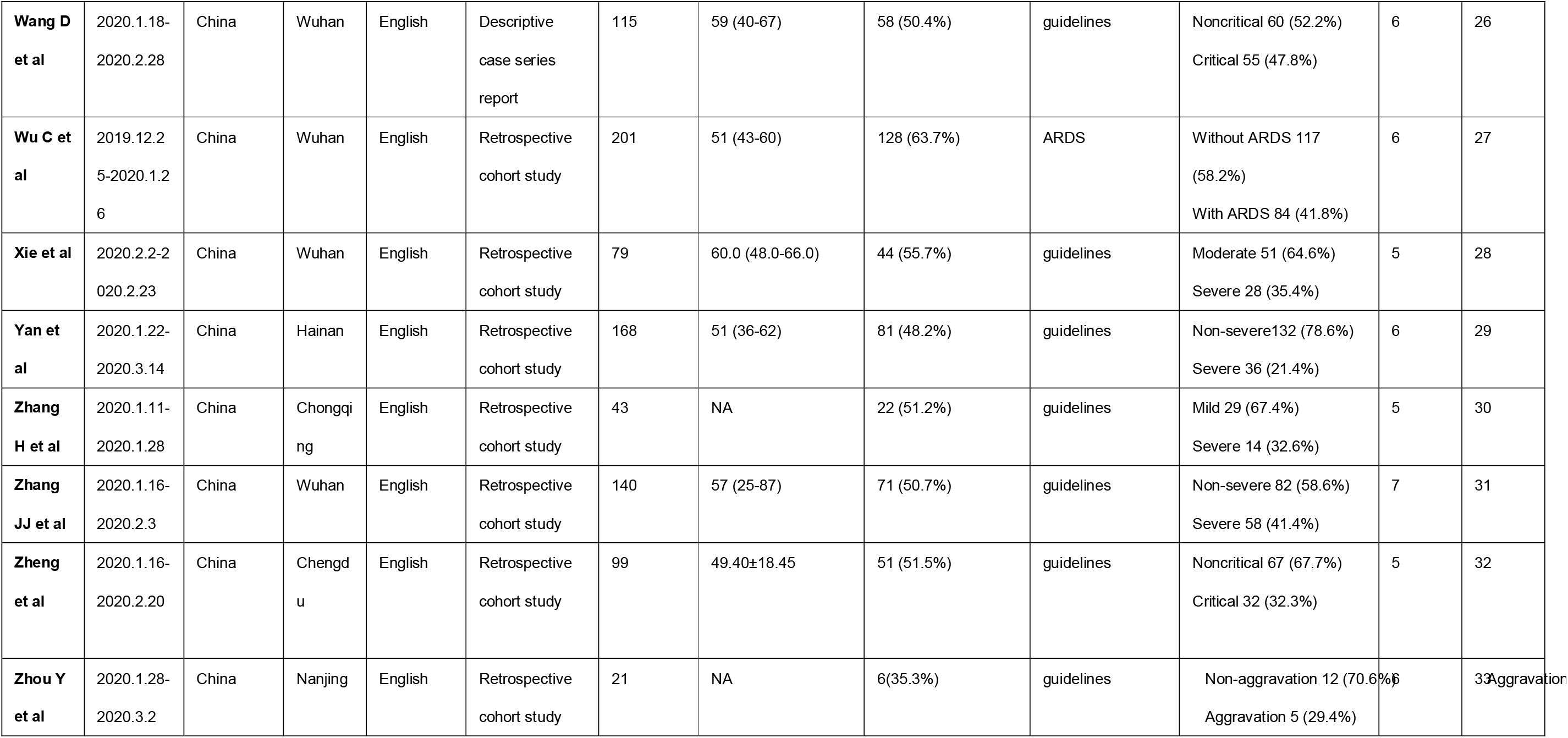
Main characteristics of the included studies in meta-analysis.

### Coagulation parameters in less severe and more severe patients

The D-dimer, PLT, PT, APTT, and FIB were available in 25, 13, 13, 9, and 5studies respectively. We use the random-effects model due to significant heterogeneity of studies on D-dimer, PLT, PT, and APTT. Notably, D-dimer and PT were significantly higher in more severe patients than in less severe patients (SMD 0.83, 95% CI: 0.70-0.97, I^2^ 56.9%; SMD 0.39, 95% CI: 0.14-0.64, I^2^ 77.9% respectively). The fixed-effects model was used due to the insignificant heterogeneity of studies on FIB. The result showed that patients with more severe pneumonia exhibited significantly higher FIB compared to less severe patients (SMD 0.35,95% CI: 0.17-0.53, I^2^ 42.4%).

However, no significant difference was observed in PLT and APTT values between severe and mild patients (SMD −0.26, 95% CI: −0.56-0.05, I2 82.2%; SMD −0.14, 95% CI:-0.45-0.18, I2 75.5% respectively). A forest plot of the coagulation parameters is shown in Figure 2 and Figure 3. Details of the meta-analysis are highlighted in Table 2.

**Table 2.**
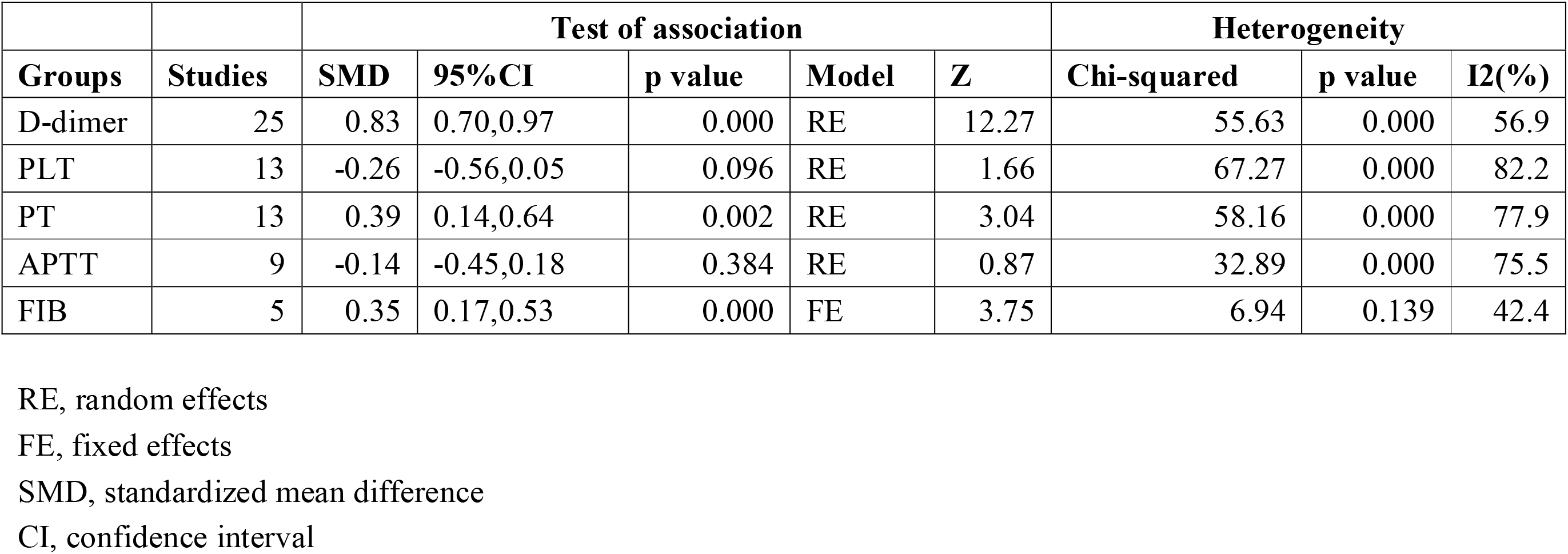
Summary of the meta-analysis results.

**Figure 2.**
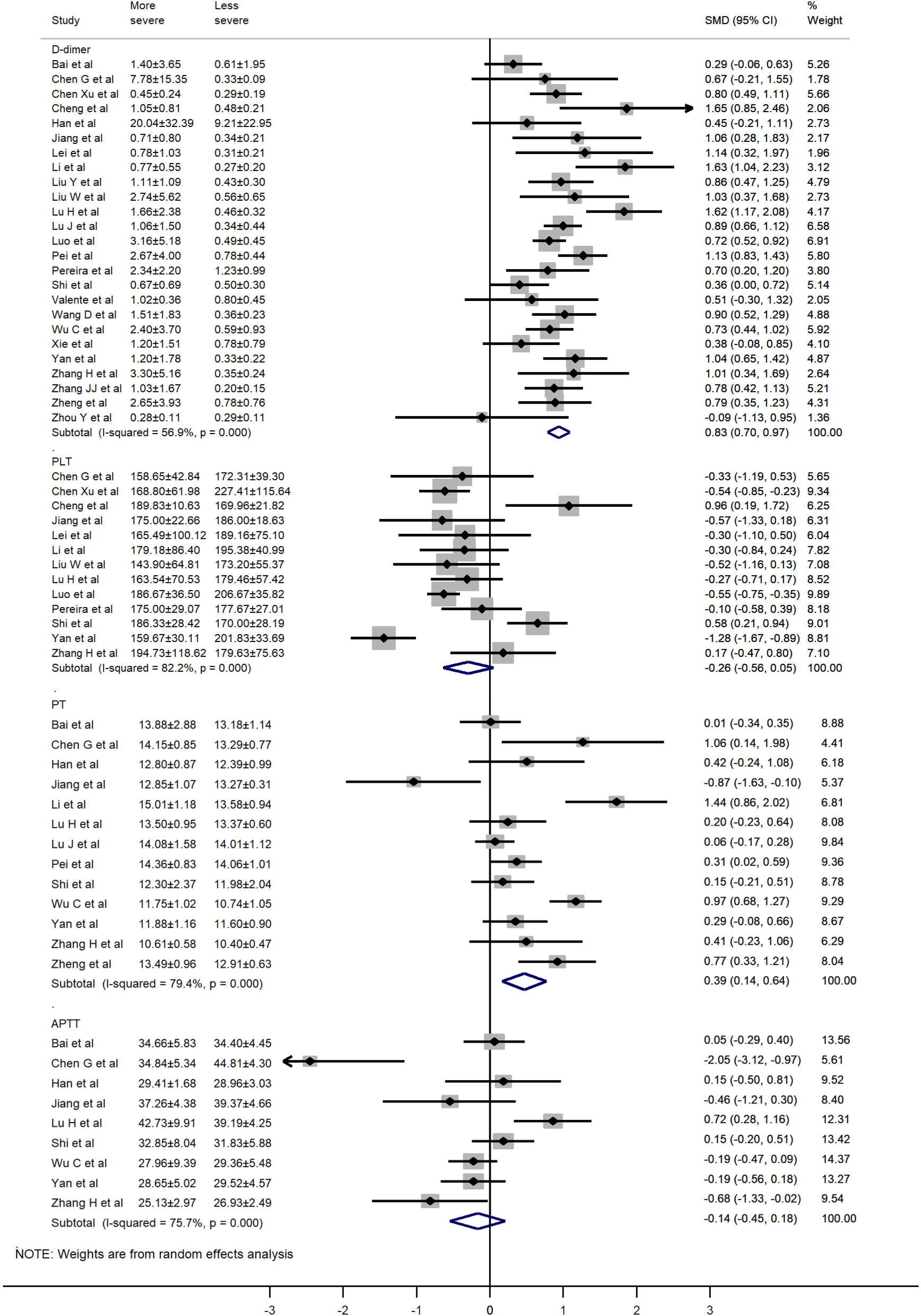
Forest plot of the association between D-dimer, PLT, PT, APTT in patients with COVID-19 stratified by disease severity.

**Figure 3.**
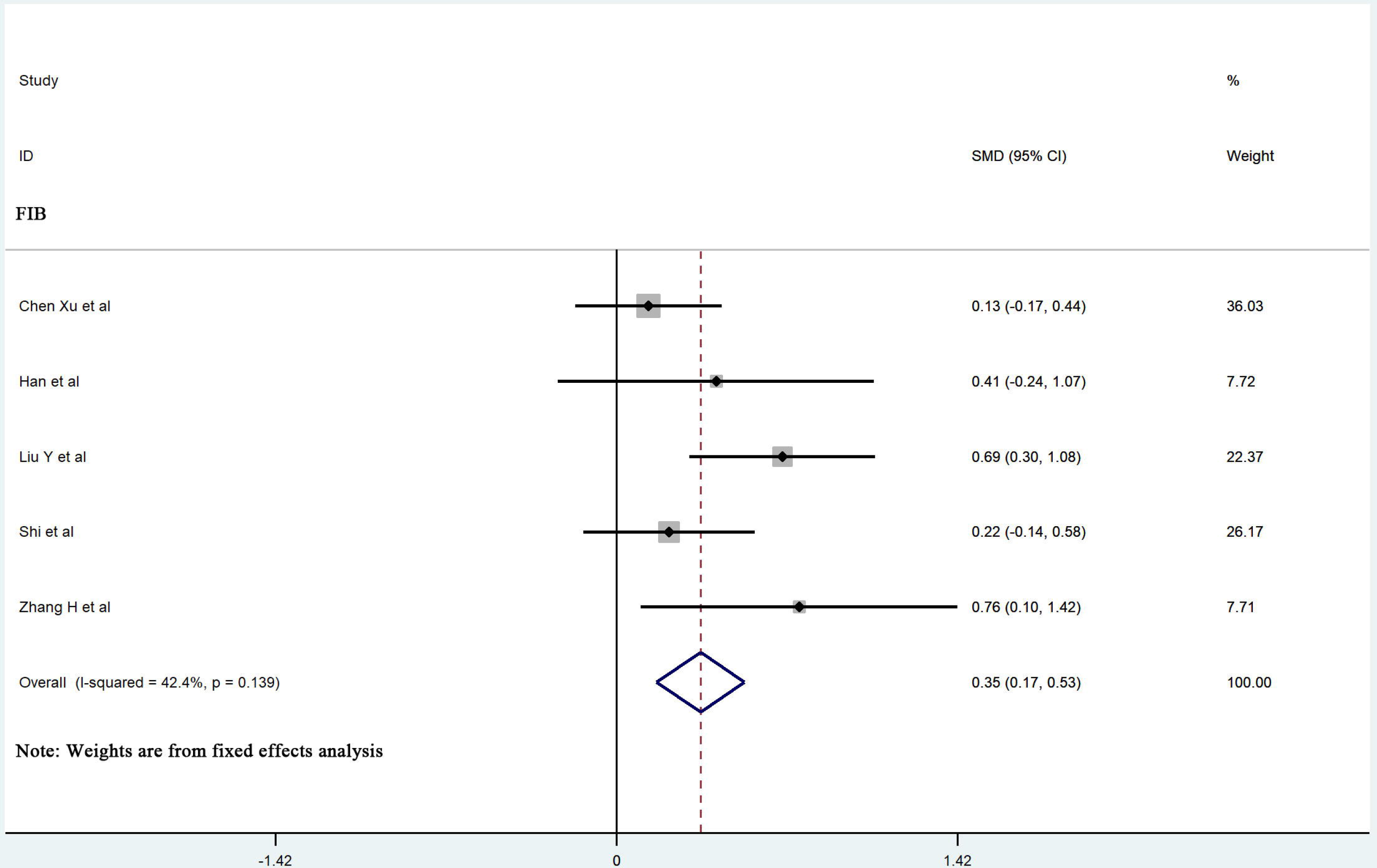
Forest plot of the association between FIB in patients with COVID-19 stratified by disease severity.

### Subgroup analysis

Further, we conducted a subgroup analysis based on location, severity criteria, and age (the median value above and below 50) to determine the sources of heterogeneity. The results for subgroup analysis are presented in Table 3. Notably, the heterogeneity was significant in the subgroup analyses. However, our subgroup analyses did not explain the observed heterogeneity of studies on D-dimer, PLT, PT, and APTT.

**Table 3.**
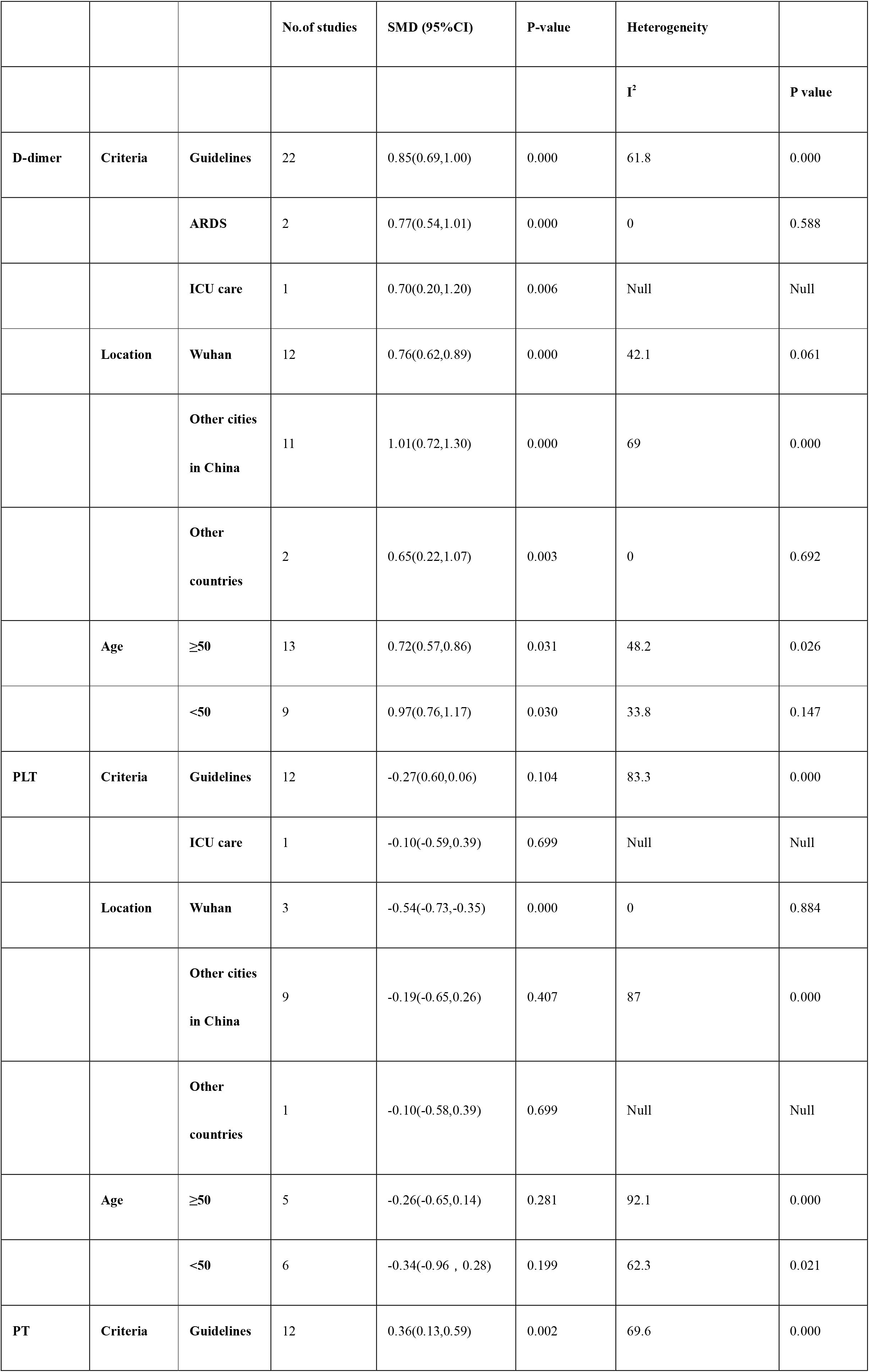
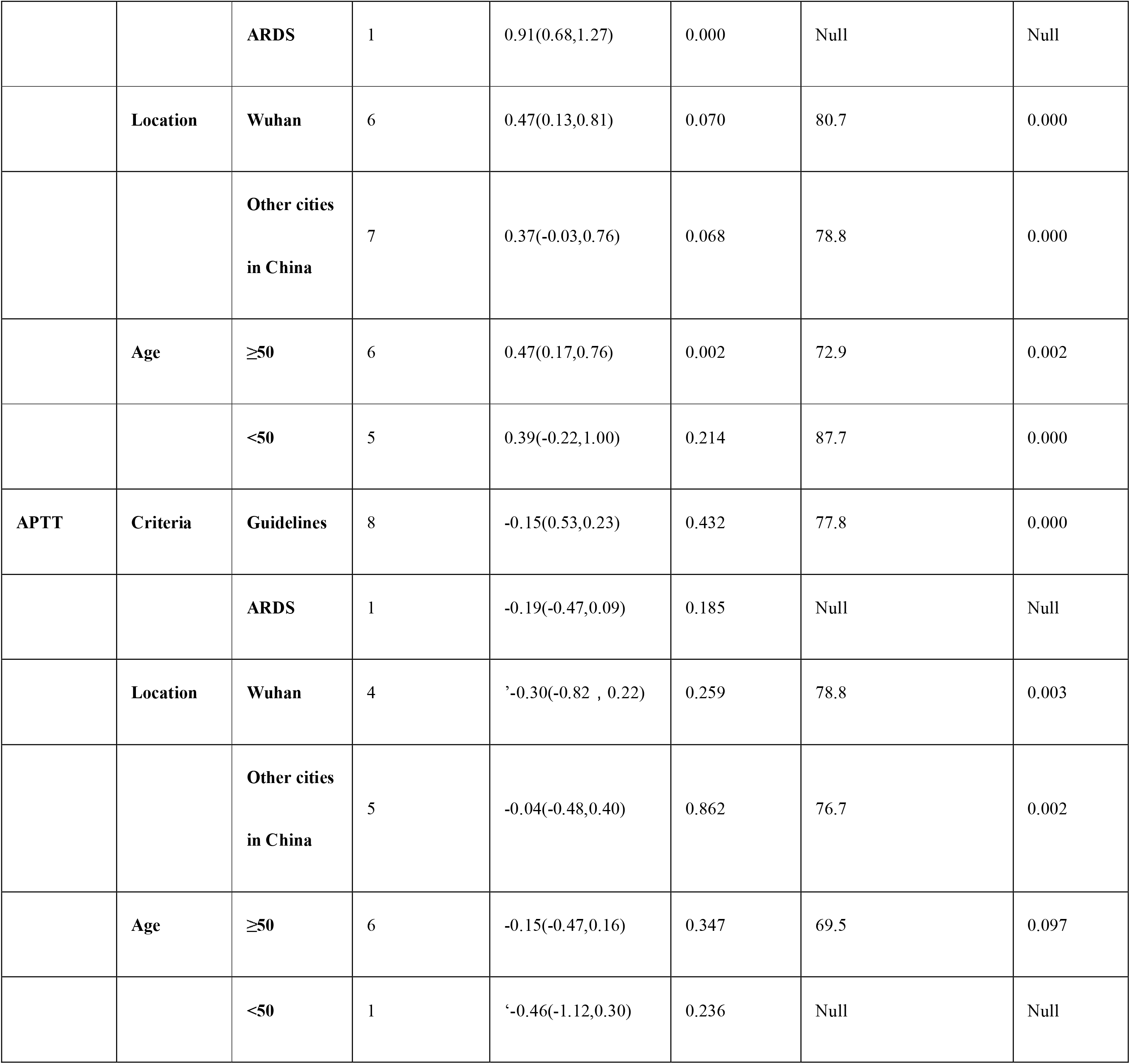
Results of subgroup meta-analyses.

### Sensitivity analysis and publication bias

Herein, sensitivity analysis showed that the pooled results were not sensitive to any individual study, this confirmed the robustness of the study findings. Detailed results of sensitivity analysis are shown in Supplementary Figure 1. Also, we assessed publication bias using the funnel plots and Egger’s regression test. There was no significant publication bias among the levels of APTT, D-dimer, FIB, PLT, PT in our study (Egger test: P=0.236, P=0.556, P=0.289, P=0.308, P=0.534 respectively). The funnel diagram and Egger’s test are described in Figure 4.

**Figure 4.**
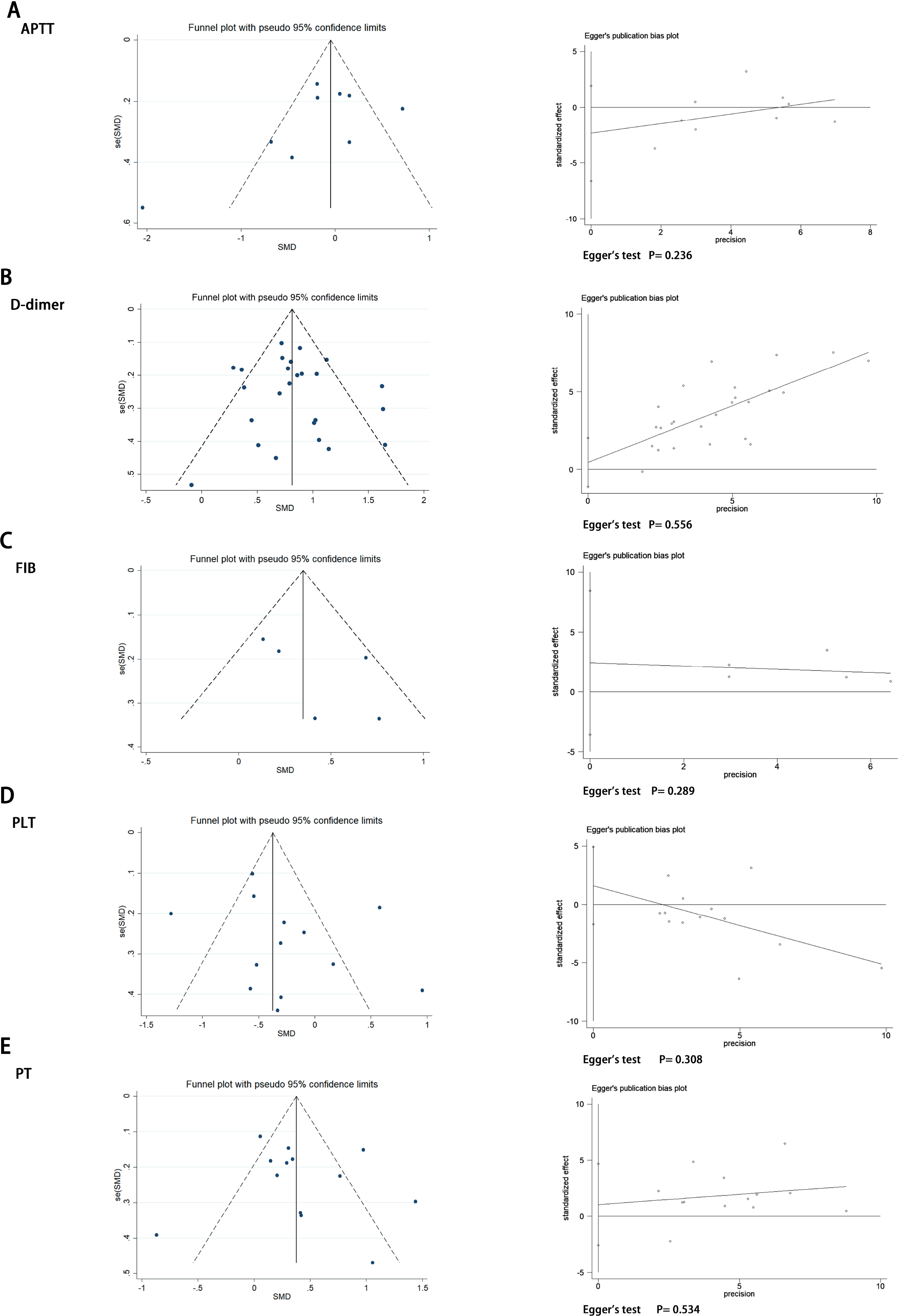
Funnel plot and Egger’s test evaluating the publication bias of (A) APTT, (B) D-dimer, (C) FIB, (D) PLT, (E) PT.

## Discussion

In this work, we revealed that coagulopathy is associated with the severity of COVID-19, and it is characterized by significantly elevated levels of D-dimer, PT, and FIB in more severe cases. This review concurs with the conclusions from the previous meta-analyses on the coagulative function of COVID-19 which found that PT and D-dimer are significantly higher in more severe patients [34]. However, we observed no significant decrease in the levels of PLT in more severe COVID-19 patients. Contrarily, a previous meta-analysis that included nine studies showed an obvious heterogeneity and may lead to unstable results [35].

Moreover, increasing levels of D-dimer and PT indicates that the fibrinolytic system is activated. The coagulation cascade is activated in viral infections as a host defense mechanism to limit the spread of pathogens. Additionally, increased production of cytokines during virus infection stimulates procoagulant reactions that highly express tissue factors, a major initiator that activates coagulation. Besides, endothelial cell activation and damage may alter the natural antithrombotic state given the tropism of the virus for ACE2 receptors [36]. The acronym COVID-19 associated coagulopathy (CAC) describes the prothrombotic properties in infected patients [37]. Although coagulopathy is reminiscent of disseminated intravascular coagulation, CAC does not cause clinical bleeding which is remarkably distinct from DIC [38]. Evidence from clinical observations shows that CAC is distinct from DIC, featured by elevated fibrinogen, and nearly the normal platelet count [38]. This observation is consistent with our findings. This can be explained by the extramedullary megakaryocytes within the microvessels that have been detected in autopsy among decedents of COVID-19 [39]. The megakaryocytes can actively produce platelets within the peripheral circulation [40].

Of note, we report the first meta-analysis that assessed the fibrinogen levels on the severity of COVID-19. Similarly, several reports showed that fibrinogen significantly increases in initial and progressive stages of virus infection. Furthermore, a recent report by Ranucci *et al* [41] indicated comprehensive coagulation analyses on ICU admission with median fibrinogen levels of 7.8 g/L and increased clot strength through thromboelastometry. During the SARS-CoV-2 infection process, a cytokine storm dominated by interleukin-6 occurs [42], this has been found majorly regulate and stimulate fibrinogen synthesis. Besides, the variation in fibrinogen levels is dependent on the infection stages encountered. Elsewhere, Tang *et al* [43] reported that fibrinogen levels decline at the late stage of COVID-19 non-survivors despite increased or normal levels on admission, this is potentially caused by sepsis-induced coagulopathy.

Despite the obscure mechanism of CAC, abnormal coagulation results including elevated D-dimer and fibrin degradation products are associated with higher COVID-19 mortality rates. Thus, monitoring the levels of coagulation parameters is immensely significant in managing COVID-19. Moreover, Zhou *et al* [5] conducted a study in 191 patients from Wuhan and found that D-dimer levels higher than 1ug/L are associated with higher in-hospital mortality. The World Health Organization [44] recently issued guidelines for managing COVID-19, whereby it suggested that great focus should be geared on coagulation dysfunction and thrombotic events.

Based on our findings, we also suggest that dynamic monitoring of coagulation parameters in hospitalized COVID-19 patients is necessary for predicting COVID-19 progression that is associated with unfavorable outcome and early thrombotic events. Moreover, futures studies should determine whether early anticoagulation features such as low molecular weight heparin (LMWH), warfarin, or new oral anticoagulant (NOAC) are beneficial in patients with significantly elevated coagulation parameters.

Besides, our meta-analysis has a few limitations. First, there was obvious heterogeneity among studies regarding definitions of the COVID-19 severity, and either subgroup or sensitivity analyses could not identify the heterogeneity source. Some selected studies did not report using mean and standard deviation, however, they gave estimates by median and quartile which may lead to deviation and effect heterogeneity. We excluded the low-quality studies to maintain the reliability of the conclusion. Second, this meta-analysis was conducted for studies that failed to describe all relevant characteristics of individual patients and it was difficult to adjust on the potentially confounding factors such as comorbidity and treatment (including use of anticoagulation or glucocorticoid). Finally, we included retrospective studies in the meta-analysis and there are risks of bias in the data collected.

In conclusion, coagulopathy is associated with the severity of COVID-19. The D-dimer, PT, and FIB are dominant parameters for evaluating coagulopathy in COVID-19. COVID-19 associated coagulopathy has different features from DIC, therefore, it is necessary to closely monitor the dynamics of coagulation parameters.

## Methods

### Search strategy

The protocol for this systematic review and meta-analysis has been registered on the International Platform of Registered Systematic Review and Meta-analysis Protocols (INPLSY) as INPLASY2020500049 (https://inplasy.com/). The reports included in the systematic review follows the Meta-analysis of Observational Studies in Epidemiology (MOOSE) guidelines (the checklist is shown in Supplementary Table 1). The literature reports were written in English and Chinese. We conducted a literature search on the electronic databases including Pubmed, Embase, Cochrane, WanFang Database, CNKI and medRxiv for reports published from December 1, 2019 to May 1, 2020 using a combination of the following keywords: “COVID-19” or “2019 novel coronavirus infection” or “SARS-CoV-2” and” characteristics” or” coagulopathy” or “coagulation”. Besides, we screened and conducted a manual search of the references listed in each article to obtain comprehensive results. The search was done independently by two authors (Xiaolin Zhang and Xue Yang). A third investigator (Hongmei Jiao) came in to resolve any contradicting search results.

### Inclusion and exclusion criteria

The inclusion criteria were as follows: (1) Original studies focused on clinical characteristics of patients with COVID-19 including observational study, case-control studies, cohort studies, and randomized control studies; (2) patients were categorized into less and more severe groups; (3) the coagulation parameters between groups were described. Exclusion criteria were as follows: (1) Non-original studies including commentaries, editorials, case reports, letters, meta-analysis, guidelines, and family-based studies; (2) same patients are enrolled in different articles; (3) patients in studies are below 18□years old.

### Data extraction and quality assessment

Two investigators (Xiaolin Zhang and Xue Yang) independently extracted data and evaluated the quality of literature reports. Contradicting decisions from the two authors were resolved by a third investigator (Hongmei Jiao) or through consensus. Data items extracted from each study included the study characteristics, demographic information, and outcomes of interest. We used the Newcastle-Ottawa scale (NOS) which included patient selection, study comparability, and three components of outcomes assessment to evaluate the quality of the original study. Low-quality articles (NOS<5) were excluded from this meta-analysis.

### Data analysis

All statistical analyses were performed via Stata 14.0 (Stata, College Station, TX, USA). For continuous variables, we calculated the standard mean difference (SMD) and the 95%CI. Heterogeneity among the studies was assessed using the Chi-squared and I^2^ tests. For studies that only reported median and range, CI or interquartile range, we estimated means and SDs as described by Wan [8]. A random-effects model was used when either P<0.05 or I^2^>50% defined significant heterogeneity across the articles. Otherwise, we used the fixed-effects model. A p-value of less than 0.05 was considered significant statistically. Further, a sensitivity analysis was conducted to evaluate the stability of the results and determine the effect of individual study on pooled results. Evidence of publication bias was examined using Egger’s regression test for funnel asymmetry in addition to the visual inspection of funnel plots.

## Data Availability

All data are fully available without restriction.

## ACKNOWLEDGMENT

None.

## CONFLICTS OF INTEREST

Xiaolin Zhang, Xue Yang, Hongmei Jiao, Xinmin Liu declare that they have no competing interests.

## AUTHOR CONTRIBUTIONS

Study conception and design: Xiaolin Zhang. Data acquisition: Xiaolin Zhang, Xue Yang. Statistical analysis: Xiaolin Zhang. Interpretation of the data: Xiaolin Zhang, Xue Yang. Drafting of the manuscript: all authors. Critical revision the manuscript for important intellectual content: all authors. Final approval of the manuscript: all authors.

## Funding information

This work was supported by the Scientific Research Seed Fund of Peking University First Hospital (Grant No. 2018SF058).

### Acronyms and Abbreviations

ACE2: Angiotensin-Converting Enzyme 2
APTT: Activated partial thromboplastin time
ARDS: Acute respiratory distress syndrome
CAC: COVID-19 associated coagulopathy
CI: Confidence interval
COVID-19: Coronavirus disease 2019
DIC: Disseminated intravascular coagulation
FIB: Fibrinogen
LMWH: Low molecular weight heparin
MERS: Middle East Respiratory Syndrome
NOAC: New oral anticoagulant
PLT: Platelet
PT: Prothrombin time
SARS: Severe Acute Respiratory Syndrome
SARS-CoV-2: Severe Acute Respiratory Syndrome Coronavirus 2
SD: Standard deviation
SMD: Standard mean difference

## Notes

### Competing Interest Statement

The authors have declared no competing interest.

### Author Declarations

The study was also approved by your Institutional Ethics Committe.

